# Association between *ALDH2* genotypes and atrial fibrillation recurrence following catheter ablation: prospective multicenter cohort study

**DOI:** 10.1101/2024.06.28.24309692

**Authors:** Tadashi Hoshiyama, Keiichi Ashikaga, Kenji Morihisa, Miwa Ito, Kentaro Oniki, Junji Saruwatari, Masanobu Ishii, Hisanori Kanazawa, Hitoshi Sumi, Shozo Kaneko, Takuya Kiyama, Yuichiro Tsuruta, Kohei Matsunaga, Yuta Tsurusaki, Kenichi Tsujita

**Affiliations:** Department of Cardiovascular Medicine, Graduate School of Medical Sciences, Kumamoto University, Kumamoto, Japan; Miyazaki Medical Association Hospital Cardiovascular Center, Miyazaki, Japan; Division of Cardiology, Kumamoto Chuo Hospital, Kumamoto, Japan; Division of Pharmacology and Therapeutics, Graduate School of Pharmaceutical Sciences, Kumamoto University, Kumamoto, Japan

**Keywords:** Atrial fibrillation, Catheter ablation, Alcohol consumption, *ALDH2* genotype, Alcohol flushing syndrome, Recurrence risk factor

## Abstract

**Background:** Alcohol, a risk factor for atrial fibrillation (AF), is metabolized by aldehyde dehydrogenase 2 (ALDH2). Notably, alcohol flushing syndrome attributed to the dysfunctional alleles of *ALDH2* (*ALDH2*-deficient variant) carriers are prevalent among East Asian populations. These patients are at risk for developing AF when accompanied with habitual alcohol consumption. However, the effect of the *ALDH2* genotype on catheter ablation, the most successful treatment option for AF, remains unclear.

**Methods:** Totally 371 patients who underwent their first catheter ablation for AF were enrolled in this prospective cohort study. They were categorized into four groups based on their *ALDH2* genotypes and habitual alcohol consumption to understand the contribution status to their impact on the risk of AF recurrence. The primary outcome was to determine the proportion of AF recurrence among the four groups during a 1-year follow-up period using Kaplan–Meier analysis. The secondary outcome involved assessing the contributions of each group to AF recurrence and other risk factors using multivariate analysis.

**Results:** This study comprised 239 *ALDH2*-wild type (147 habitual drinkers) and 132 *ALDH2*-deficient variant carriers (31 habitual drinkers). Kaplan–Meier curves indicated that *ALDH2*-deficient variant carriers with habitual alcohol consumption exhibited the highest proportion of AF recurrence compared with the other groups (p<0.01). In addition, ALDH2-deficient variant itself was not associated with AF recurrence (hazard ratio [HR]=1.56, p=0.10), *ALDH2*-deficient variant carriers with habitual alcohol consumption exhibited a higher HR (HR=5.01, p=0.02) in multivariate analysis. Notably, it conferred a higher risk than that for ALDH2 wild-type patients with habitual alcohol consumption (HR=2.36, p=0.02).

**Conclusion:** While the *ALDH2*-deficient variant itself showed no correlation with AF recurrence, it emerged as a significant risk factor for AF when accompanied with habitual alcohol consumption. Thus, abstinence from alcohol may be necessary, even after catheter ablation is performed, especially for patients with the *ALDH2*-deficient variant.

**Clinical Perspective:** *What is Known?:*

- Alcohol, a risk factor for atrial fibrillation (AF), is metabolized by aldehyde dehydrogenase 2 (ALDH2); notably, alcohol flushing syndrome owing to dysfunctional alleles of *ALDH2* (*ALDH2*-deficient variant) is prevalent among East Asians.
- However, the relationship between *ALDH2* genotypes and AF recurrence following catheter ablation has not been clarified yet.

*What the Study Adds:*

- While the *ALDH2*-deficient variant itself was not associated with AF recurrence, it emerged as a major risk factor for AF recurrence when accompanied with habitual alcohol consumption.
- Abstinence from alcohol consumption may be necessary, even after catheter ablation, especially for *ALDH2*-deficient variant carriers.

## Introduction

Atrial fibrillation (AF) is the most common arrhythmia detected in clinical practice (1,2). This condition elevates the risks of stroke, cardiovascular diseases, and all-cause mortality (1,3). To mitigate these risks, treatment such as antiarrhythmic drug (AAD) therapy and catheter ablation, which electrically isolates the pulmonary veins (PVs), (the source of AF (4)), have been employed. In particular, catheter ablation has emerged as the preferred first-line therapy over the years (5) owing to improved clinical outcomes, as reported in a meta-analysis (6). However, the rate of recurrence within a 1-year follow-up period remains suboptimal, ranging between 10 and 40 % (7–10). Notably, non-PV foci originating outside the PVs, have been identified as a major cause of AF recurrence, posing a substantial challenge (11,12).

This non-PV foci is closely related to the AF substrate representing the low-voltage area in the atrium (13). Among various risk factors for AF substrates, including obesity (14), valve disease (15), and cardiomyopathy (16), recent attention has focused on habitual alcohol consumption (17). Studies have indicated that there is a progressive widening in atrium with increased alcohol consumption (18). And, habitual alcohol consumption is reported to be associated with AF recurrence following catheter ablation (19), promoting guidelines to recommend reducing alcohol intake, even if catheter ablation has been performed (1).

In contrast, alcohol is primary metabolized by aldehyde dehydrogenase 2 (ALDH2) (20). The *ALDH2* gene exhibits a genetic polymorphism comprising *ALDH2* homozygous wild-type (*1/*1), *ALDH2* heterozygous-deficient allele (*1/*2), and *ALDH2* homozygous-deficient allele (*2/*2). Among them, *ALDH2*1/*2* and *ALDH2*2/*2* carriers are considered as *ALDH2*-deficient variant carriers owing to the reduced or negligible activity of ALDH2, and they experience alcohol flushing syndromes such as facial flushing, nausea, palpitation, and headache after alcohol consumption (21). Notably, the proportion of these carriers is reported to be as high as 30 - 40 % among East Asian populations (22,23). Regarding the relationship between *ALDH2* genotype and AF, it has previously been reported that *ALDH2*-deficient variant carriers themselves are not more prone to AF. However, when accompanied with habitual alcohol consumption, *ALDH2*-deficient variant carriers have a heightened risk for developing AF compared with individuals having other identified risk factors (24). These findings imply that there is an increased prevalence of AF recurrence even if catheter ablation is performed in *ALDH2*-deficient variant carriers who habitually consume alcohol owing to low metabolic activity of ALDH2.

Despite this, the effect of *ALDH2* genotypes on AF recurrence following catheter ablation remains unclear. This study aimed to investigate the contribution of *ALDH2* genotypes on AF recurrence following catheter ablation, including comparisons with other risk factors.

## Methods

### Patient population

Between 2020 and 2022, a total of 373 patients who underwent their first catheter ablation for AF at three medical centers in Japan were enrolled in this multicenter prospective cohort study. The following patients were excluded: those with a history of prior catheter ablation for AF; those who underwent the Maze procedure; those with severe valve disease. Additionally, as the proportion of *ALDH2*-deficient variant carriers is higher among East-Asian population, patients from other populations were also excluded. This study was approved by the Human Research Committee of each participating institution (ethical approval number; genome 466), and written informed consent was obtained from all enrolled patients.

### Study protocol

The *ALDH2* genotypes were analyzed in all enrolled patients. These genotypes were divided into *ALDH2* homozygous wild-type (*1/*1) and *ALDH2*-deficient variant carriers, including the heterozygous-deficient (*1/*2) and homozygous-deficient (*2/*2) alleles. The genotypes of each patient were not informed until the end of the follow-up period. Moreover, the operators were unaware of the patients’ genotypes before 12 months of the follow-up period.

Following catheter ablation for AF, the patients were categorized into four groups based on their *ALDH2* genotypes and habitual alcohol consumption status. The groups were as follows: *ALDH2* wild-type carriers who did not habitually consume alcohol, *ALDH2* wild-type carriers who habitually consumed alcohol, *ALDH2*-deficient variant carriers who did not habitually consume alcohol, and *ALDH2*-deficient variant carriers who habitually consumed alcohol. It is important to note that no guidance on alcohol abstinence was provided during this study period. Under these circumstances, the primary assessment was the comparison of the AF recurrence rate among the four categories during a 1-year follow-up period using the Kaplan–Meier analysis methods. Additionally, the secondary evaluation of this study involved assessing the impact of each of the four categories on AF recurrence, considering other risk factors for AF recurrence through multivariate analysis.

### Catheter ablation

The purpose of catheter ablation was to achieve and maintaining the sinus rhythm before completing the procedure. During the procedure, circumferential PV isolation was initially performed using an irrigation catheter (ThermoCool Smart Touch SF, Biosense Webster, Irvine, CA, USA) using the CARTO 3D mapping system (Biosense Webster). Superior vena cava (SVC) isolation, cavotricuspid isthmus linear ablation, atrial linear ablation, and non-PV foci ablation were performed at the discretion of the operator. Before concluding the procedure, a left atrial voltage map was created to identify the low-voltage area which represents the AF substrate using a multipolar mapping catheter (PENTARAY catheter, Biosense Webster) under the sinus rhythm or coronary sinus pacing rhythm. The percentage of the low-voltage area in the left atrium was measured based on the definition of low-voltage area defined as bipolar voltage below 0.5 mV.

### Follow up

Discontinuation of class I or III AADs were encouraged during the 3-months blanking period after catheter ablation. Surface electrograms (ECG) was performed at 1-, 3-, 6-, and 12-months post-procedure during outpatient clinic visitations. Additionally, the 1-week Holter ECG was performed at 6- and 12-months post procedure. For patients with implanted cardiovascular implantable electronic devices, AF recurrence was evaluated through device interrogation. AF recurrence was defined as the presence of AF or atrial tachycardia lasting over 30 seconds after the 3-months blanking period.

### Identification of risk factors (variables) for AF recurrence using multivariate analysis

In addition to the *ALDH2* genotype, hypertension, obesity, sex, cardiomyopathy, valve diseases, large left atrium, and non-paroxysmal AF have been identified as risk factors for AF recurrence according to previous reports (14–16,25–27). Hypertension was defined in patients as a blood pressure of >140/90 mmHg or administration of antihypertensive medication. Obesity was characterized by a body mass index (BMI) of >30 kg/m^2^. Cardiomyopathies included dilated cardiomyopathy, hypertrophic cardiomyopathy, ischemic heart disease, and cardiac amyloidosis. Valve diseases included moderate aortic and mitral regurgitation. A large left atrium was defined as having a left atrial diameter of over 45 mm which measured by echocardiography. Non-paroxysmal AF was defined as AF lasting more than 7 days or AF requiring cardioversion for termination.

### Habitual alcohol consumption and average alcohol intake volume

Patients’ habitual alcohol consumption status was confirmed before performing catheter ablation for AF. The alcohol consumption data of each patient were obtained based on their drinking habits spanning from months to years. Habitual alcohol consumption and average alcohol intake volume were defined as previously described (17), (24). Habitual alcohol consumption was defined as >3 drinking opportunities per week, according to the Ministry of Health, Labour and Welfare in Japan. In addition, the average alcohol intake volume was represented in standard drinks per week. One standard drink was defined as 12 g of alcohol. According to this definition, alcohol intake volume was divided into the following categories: non-drinkers: 0-1 drink/week; mild-drinkers: 2-7 drinks/week; moderate drinkers: 8-21 drinks/week; and heavy drinkers: >22 drinks/week (17,24).

### *ALDH2* genotype and allele ratio

Genomic DNA used for genotyping of *ALDH2* was extracted from whole blood using a DNA purification kit (Flex Gene DNA kit, Qiagen). The *ALDH2* rs671 (Glu504Lys; *ALDH2*2*) genotypes were determined by performing a real-time TaqMan allelic discrimination assay (Step One Plus Real-Time PCR system version 2.1, Applied Biosystems) according to the manufacturer’s protocol (assay no. C_11703892_10). The allele ratio was analyzed using this method.

### Sample size calculations

The prevalence of *ALDH2*-deficient variant carriers was assumed to be within the range of 30 – 40% among the East-Asian population (21,24). And, based on catheter ablation results in both East-Asian and Caucasian populations for the patient with and without habitual alcohol consumption, the median AF free survival was found to be 13 months for *ALDH2* wild-type carriers and 7 months for *ALDH2*-deficient variant carriers with habitual alcohol consumption (7), (10). In addition, in report on the relationship between *ALDH2* genotypes and AF development, the proportions of *ALDH2* wild-type and *ALDH2*-deficient variant carriers with habitual alcohol consumption were 35 and 8 %, respectively (24). Consequently, 30 *ALDH2*-deficient variant and 135 *ALDH2* wild-type carriers with habitual alcohol consumption were calculated to be needed with a significance level of 0.05 (type I error probability) and a power of 0.8. Consequently to evaluate AF free survival rates using Kaplan–Meier analysis, a minimum of 360 patients was deemed necessary. Similarly, at least 288 patients were required to evaluate the hazard ratio (HR) for the AF-free survival rate in *ALDH2* wild-type and *ALDH2*-deficient variant carriers with habitual alcohol consumption.

### Statistical analysis

All continuous data exhibited a skewed distribution except for the left atrial diameter, as confirmed by the Shapiro–Wilk test. Therefore, continuous data, except the left atrial diameter were expressed as the medians (interquartile range) and analyzed using non-parametric tests. Differences among multiple groups were assessed using the Kruskal– Wallis test. The left atrial diameter was expressed as the mean ± standard deviation and analyzed using parametric tests. Categorical data were presented as numbers or percentages. Kaplan–Meier analysis was conducted with the log-rank test. Following the Kaplan–Meier analysis, a post hoc analysis was performed using the holm test.

Multivariate analyses were performed using a Cox proportional hazard model with HR and their 95% confidence intervals (CI). A P value < 0.05 was considered to indicate statistical significance. Statistical analyses were performed using R software (R foundation for Statistical Computing, Vienna, Austria).

## Results

### Patient characteristics

Among the 373 patients enrolled, two were excluded from the study owing to death during the 3-month blanking period. Additionally, three patients were lost to follow-up, and one died during follow-up, these events occurred after the 3-months blanking period. Therefore, these patients were included in evaluation until the final follow-up data. Consequently, 371 patients were ultimately included in this study.

Within this cohort, the numbers of *ALDH2*1/*1* (*ALDH2* wild-type) and *ALDH2*-deficient variant carriers, including *ALDH2*1/*2* and *ALDH2*2/*2*, were 239 and 132 (115 and 17), respectively. The distribution of *ALDH2* genotypes in this study was consistent with previous reports in East-Asians, including the Japanese population (21), (24).

Table 1 summarizes the demographic and clinical –characteristics of the patients’ –. Notably, there were no significant differences between *ALDH2* wild-type and *ALDH2*-deficient variant carriers, except for habitual alcohol consumption. The prevalence of habitual alcohol consumption in *ALDH2*-deficient variant carriers was significantly lower than that in *ALDH2* wild-type carriers (31/132 patients, 23.5 % vs. 147/239 patients, 64.5 %; p < 0.01). Binge drinking was not observed among these patients. Furthermore, with respect to the proportion of AF types and procedural characteristics within each *ALDH2* genotype, no significant differences were observed between *ALDH2* wild-type and *ALDH2*-deficient variant carriers.

**Table 1.** Patients’ characteristics.

|  | All<br>n=371 | ALDH2-wild<br>n=239 | ALDH2 variant<br>n=132 | p value |
| --- | --- | --- | --- | --- |
| **Background characteristics** |  |  |  |  |
| Age (years) | 70.0 (62.5–75.0) | 69.0 (62.5–74.0) | 71.0 (62.8–76.3) | 0.07 |
| Male: n (%) | 246 (66.3) | 156 (65.3) | 90 (68.2) | 0.65 |
| BMI (kg/m<sup>2</sup>) | 24.1 (22.0–26.6) | 24.1 (22.1–26.5) | 23.9 (21.5–26.8) | 0.66 |
| Habitual alcohol consumption: n (%) | 178 (48.0) | 147 (61.5) | 31 (23.5) | <0.01 |
| HT: n (%) | 236 (63.6) | 153 (64.0) | 83 (62.9) | 0.82 |
| DM: n (%) | 81 (21.8) | 51 (21.3) | 30 (22.7) | 0.79 |
| Valve disease: n (%) | 27 (7.3) | 14 (5.9) | 13 (9.8) | 0.21 |
| Cardiomyopathy: n (%) | 41 (11.1) | 22 (9.2) | 19 (14.4) | 0.17 |
| eGFR(mL/min/1.73m<sup>2</sup>) | 63.0 (53.4–74.9) | 64.0 (54.0–75.0) | 62.0 (50.0–73.3) | 0.13 |
| BNP (pg/mL) | 60.8 (21.6–125.7) | 60.8 (21.2–122.0) | 62.1 (22.0–137.8) | 0.65 |
| EF (%) | 60.6 (54.2–65.4) | 60.6 (54.7–64.9) | 60.6 (52.5–65.9) | 0.78 |
| LAD (mm) | 40.0±7.0 | 39.8±6.9 | 40.2±7.3 | 0.64 |
| **type of AF** |  |  |  | 0.74 |
| Paroxysmal: n (%) | 216 (58.2) | 141 (59.0) | 75 (56.8) |  |
| Non-paroxysmal: n (%) | 155 (41.8) | 98 (41.0) | 57 (43.2) |  |
| **Procedural characteristics** |  |  |  |  |
| Pulmonary vein isolation: n (%) | 371 (100) | 239 (100) | 132 (100) | N.A |
| SVC isolation: n (%) | 98 (26.4) | 57 (23.8) | 41 (31.1) | 0.14 |
| CTI ablation: n (%) | 121 (32.6) | 73 (30.5) | 48 (36.4) | 0.30 |
| Non-PV foci ablation: n (%) | 35 (9.4) | 19 (7.9) | 16 (12.1) | 0.20 |
| Linear ablation: n (%) | 21 (5.7) | 17 (7.1) | 4 (3.0) | 0.16 |
| Other procedure: n (%) | 12 (3.2) | 10 (4.2) | 2 (1.5) | 0.23 |

- 2 - 3 AF=atrial fibrillation; BMI=body mass index; BNP=brain natriuretic peptide; CTI=cavotricuspid isthmus; DM=diabetes mellitus; - 4 EF=ejection fraction; eGFR=estimated glomerular filtration rate; LAD=left atrial dimension; PV=pulmonary vein; SVC=superior vena - 5 cava.

### Relationship between *ALDH2* genotypes and AF recurrence

Among the 371 patients, AF recurrence was observed in 73 patients during the 1-year follow-up period. Figure 1A illustrates the Kaplan–Meier curves of AF recurrence-free survival rates, classified by *ALDH2* genotypes. As shown in Figure 1A, no significant differences in AF recurrence were found between *ALDH2*-deficient variant and *ALDH2* wild-type carriers (p = 0.97). Conversely, Figure 1B indicates a significantly higher proportion of AF recurrence in patients with habitual alcohol consumption than in patients without habitual alcohol consumption (p < 0.01).

**Figure 1.**
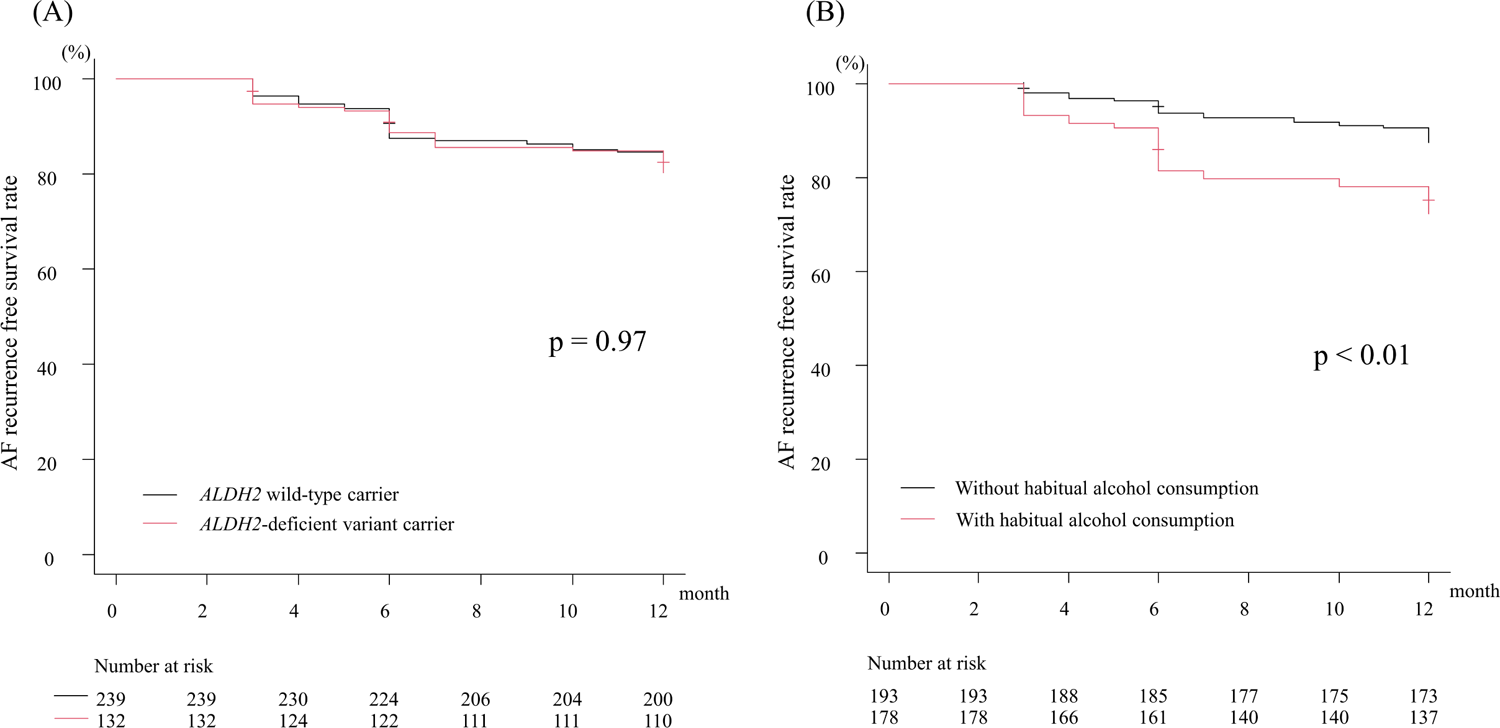
Kaplan–Meier curves illustrating AF recurrence in different *ALDH2* genotypes (panel A) and comparing AF recurrence in patients with and without habitual alcohol consumption (panel B). AF, atrial fibrillation; ALDH2, aldehyde dehydrogenase 2.

Furthermore, the multivariate analysis using a Cox proportion hazards model revealed a close relationship between habitual alcohol consumption and AF recurrence (HR = 3.41, 95% CI 1.89-6.14, p < 0.01). However, *ALDH2*-deficient variant carriers showed no association with AF recurrence in both univariate (HR = 1.01, 95% CI 0.63-1.63, p = 0.97) and multivariate analyses (HR = 1.56, 95% CI 0.91-2.67, p = 0.10) (Table 2). While, the relationship between AF recurrence and *ALDH2* genotypes in regard to habitual alcohol consumption was not analyzed in this analysis. Therefore, as previously mentioned in the methods section, the enrolled patients were divided into four groups based on their *ALDH2* genotypes and habitual alcohol consumption status. And rate of AF recurrence on each group were analyzed.

**Table 2.** Relationship between AF recurrence and variables.

|  | Univariate analysis |  |  |  | Multivariate analysis |  |  |
| --- | --- | --- | --- | --- | --- | --- | --- |
|  | HR | 95% CI | p value |  | HR | 95% CI | p value |
| HT | 0.83 | 0.52–1.33 | 0.44 |  | 0.78 | 0.49–1.26 | 0.31 |
| Obesity | 1.64 | 0.84–3.19 | 0.15 |  | 1.48 | 0.73–3.01 | 0.28 |
| Male | 1.11 | 0.70–1.82 | 0.67 |  | 0.63 | 0.36–1.09 | 0.10 |
| Cardiomyopathy | 1.02 | 0.49–2.12 | 0.96 |  | 1.11 | 0.52–2.37 | 0.78 |
| Valve disease | 1.70 | 0.81–3.54 | 0.16 |  | 1.93 | 0.87–4.00 | 0.11 |
| Large left atrium | 1.70 | 1.08–3.01 | 0.04 |  | 1.35 | 0.75–2.42 | 0.32 |
| Non-paroxysmal AF | 1.71 | 1.01–2.87 | 0.02 |  | 1.30 | 0.78–2.16 | 0.35 |
| Habitual alcohol consumption | 2.40 | 1.47–3.91 | <0.01 |  | 3.41 | 1.89–6.14 | <0.01 |
| *ALDH2*-deficient variant | 1.01 | 0.63–1.63 | 0.97 |  | 1.56 | 0.91–2.67 | 0.10 |

AF=atrial fibrillation; ALDH2= aldehyde dehydrogenase 2; HT=hypertension.

Figure 2 shows the Kaplan–Meier curves depicting AF recurrence–free survival rates categorized by the *ALDH2* genotypes and habitual alcohol consumption status. As shown in this figure, significant difference were observed among the four groups (p < 0.01). Notably, *ALDH2*-deficient variant carriers with habitual alcohol consumption exhibited a higher rate of AF recurrence compared to *ALDH2* wild-type carriers without habitual alcohol consumption (p < 0.001), *ALDH2*-deficient variant carriers without habitual alcohol consumption (p < 0.001), and *ALDH2* wild-type carriers with habitual alcohol consumption (p < 0.04). In the multivariate analysis, while it was found that *ALDH2* wild-type carriers with habitual alcohol consumption showed a close relationship with AF recurrence (HR = 2.36, 95% CI 1.12-4.95, p = 0.02), *ALDH2*-deficient variant carriers with habitual alcohol consumption had the highest level of AF recurrence (HR = 5.01, 95% CI 2.13-11.80, p < 0.01), even after accounting for other AF recurrence risk factors (Table 3). In contrast, *ALDH2* wild-type and deficient variant carriers without habitual alcohol consumption did not show any significant association with AF recurrence in both univariate and multivariate analyses. Regarding other risk factors of AF recurrence, while a large left atrium and non-paroxysmal AF demonstrated a relationship with AF recurrence in the univariate analysis (HR = 1.70, 95% CI 1.08-3.01, p = 0.04; HR = 1.71, 95% CI 1.01-2.87 p = 0.02), this relationship was not evident in the multivariate analysis (Table 3).

**Figure 2.**
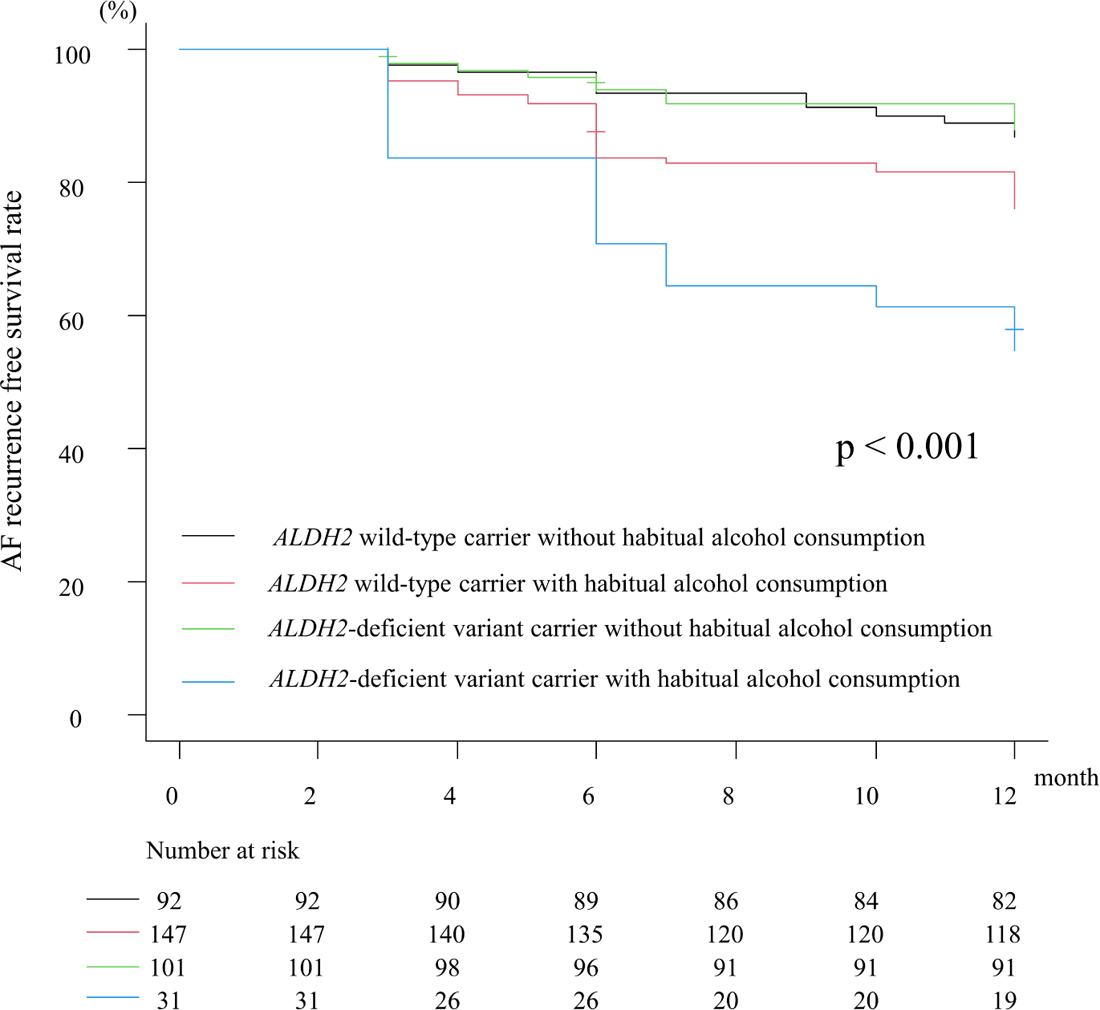
Kaplan–Meier curves depicting AF recurrence in four groups categorized by both *ALDH2* genotypes and habitual alcohol consumption status. AF, atrial fibrillation; ALDH2, aldehyde dehydrogenase 2.

**Table 3.**
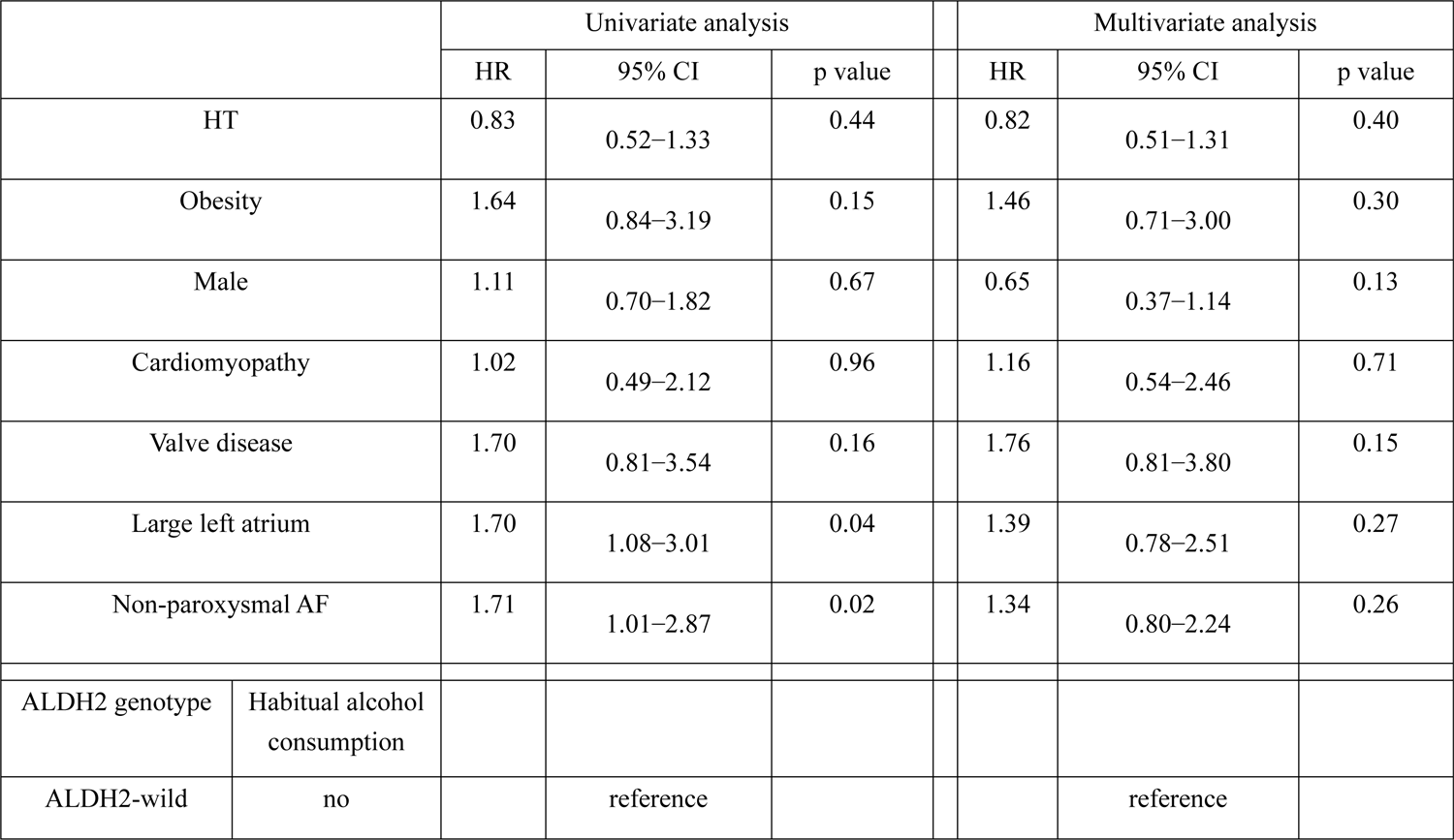
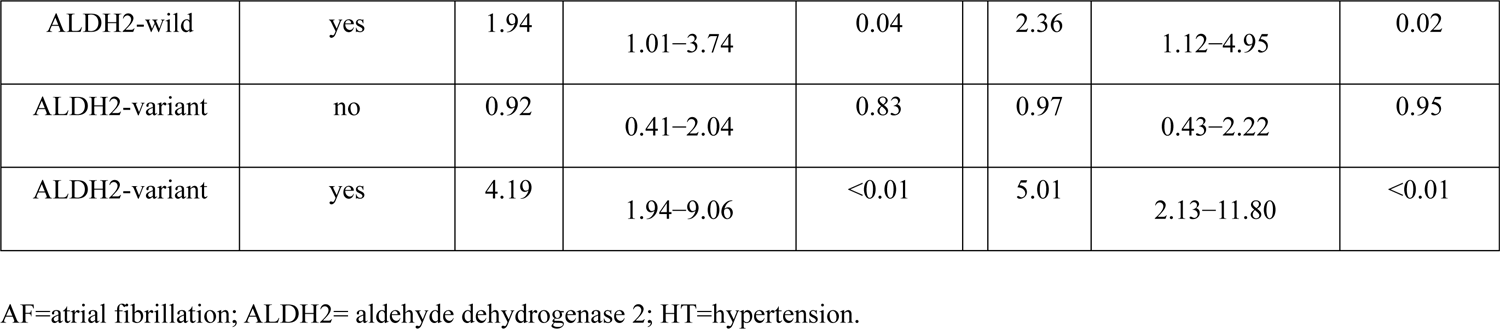
Relationship between AF recurrence and variables including groups of *ALDH2* genotypes and habitual alcohol consumption.

Furthermore, we analyzed the low-voltage area in the left atrium, which is indicative of the AF substrate, in 350 patients (a voltage map was not created for the other 21 patients). Figure 3 represents the low-voltage area of each group categorized by *ALDH2* genotypes and habitual alcohol consumption status. The percentages of low-voltage areas for *ALDH2* wild-type carriers without habitual alcohol consumption, *ALDH2* wild-type carriers with habitual alcohol consumption, *ALDH2*-deficient variant carriers without habitual alcohol consumption, and *ALDH2*-deficient variant carriers with habitual alcohol consumption were 0.10% (0 - 0.90%), 0.90% (0 - 2.50%), 0.4% (0 - 1.2%), and 1.5% (0 - 4.50%), respectively. As shown in this figure, significant difference were observed among the four groups (p < 0.01). However, while the low-voltage area in the left atrium of patients with habitual alcohol consumption was larger than that of those without habitual alcohol consumption, the low-voltage area of *ALDH2*-deficient variant carriers and *ALDH2* wild-type carriers with habitual alcohol consumption dis not significantly differ.

**Figure 3.**
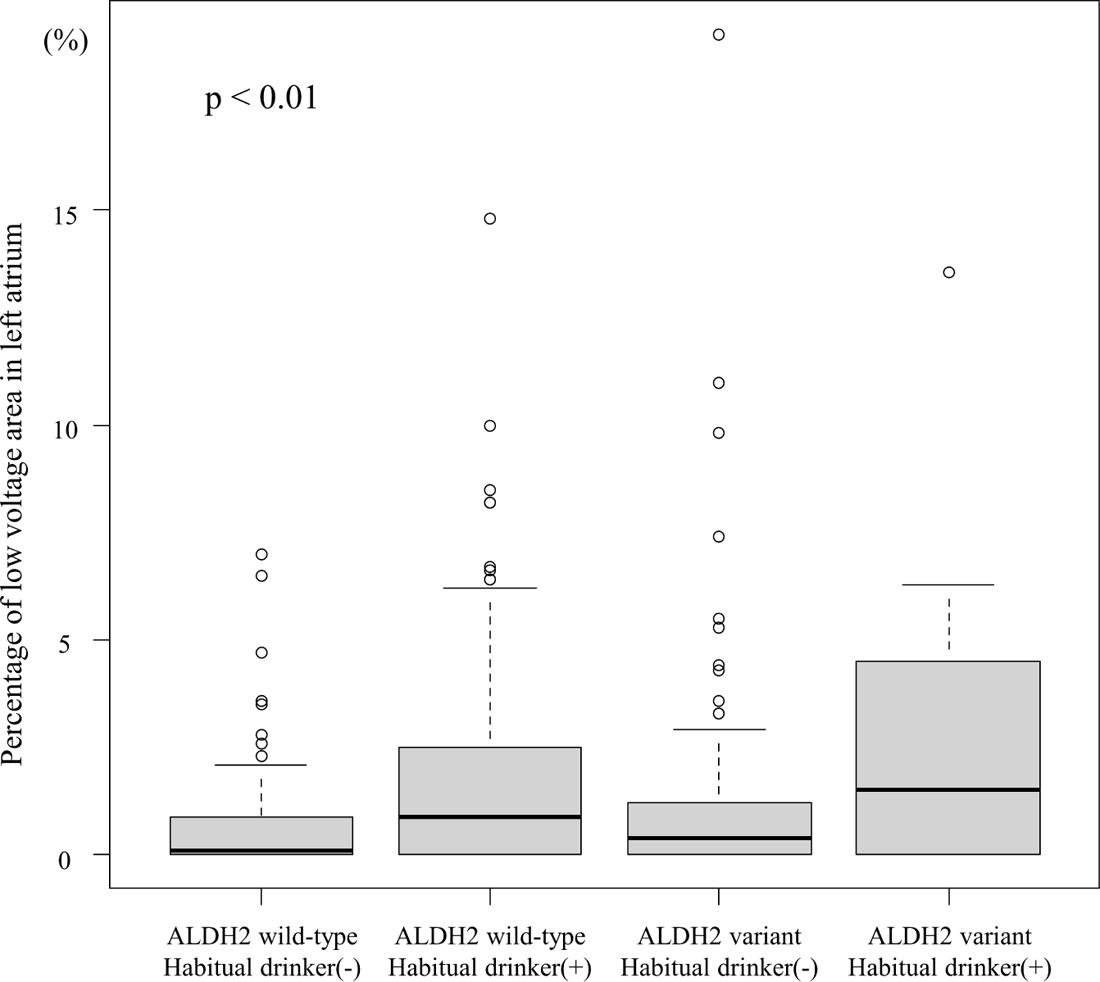
Percentage of low-voltage areas in the left atrium within the four groups classified by ALDH2 genotypes and habitual alcohol consumption status. ALDH2, aldehyde dehydrogenase 2.

### Correlation between volume of alcohol intake and prevalence of AF recurrence among *ALDH2* wild-type and *ALDH2*-deficient variant carriers

Table 4 presents the relationship between AF recurrence and alcohol intake volume categories in *ALDH2* wild-type and *ALDH2*-deficient variant carriers. AF recurrence did not exhibit a dependency on the amount of alcohol consumed by *ALDH2* wild-type carriers. Nevertheless, there was a proportional increase in the HR for AF recurrence with the amount of alcohol consumed by *ALDH2*-deficient variant carriers.

**Table 4.** Relationship between AF recurrence and alcohol consumption in each genotype.

|  | n | AF recurrence | HR | 95% CI | p value |
| --- | --- | --- | --- | --- | --- |
| ALDH2-wild type |  |  |  |  |  |
| Non-drinker | 92 | 12 | reference |  |  |
| Mild-drinker | 6 | 0 | NA | NA | NA |
| Moderate-drinker | 71 | 19 | 2.24 | 1.09–4.62 | 0.03 |
| Heavy drinker | 70 | 16 | 1.83 | 0.88–3.88 | 0.12 |
| ALDH2-variant |  |  |  |  |  |
| Non-drinker | 101 | 12 | reference |  |  |
| Mild drinker | 3 | 1 | 3.56 | 0.46–27.40 | 0.22 |
| Moderate-drinker | 19 | 8 | 4.12 | 1.68–10.10 | <0.01 |
| Heavy drinker | 9 | 5 | 5.91 | 2.08–16.81 | <0.01 |

AF=atrial fibrillation; ALDH2= aldehyde dehydrogenase 2

Regarding the volume of alcohol consumption, despite the absence of specific alcohol abstinence guidance, 13 *ALDH2* wild-type and three *ALDH2*-deficient variant carriers independently discontinued their drinking habits or reduced their alcohol consumption during the 1-year follow-up period. While the number of patients in this subgroup was small, it did not have a significant impact on the results of the statistical analysis. In contrast, the habitual alcohol consumption 358 patients’ remained unchanged.

### PV reconnection for each *ALDH2* genotypes among those who underwent a second procedure

As previously mentioned, AF recurrence occurred in 73 out of 371 patients during the 1-year follow-up period. Of these, a second procedure was performed in 32 of these patients. Among them, PV electrical reconnection was observed in two out of five (40.0 %) *ALDH2* wild-type carriers without habitual alcohol consumption, six out of 16 (37.5 %) *ALDH2* wild-type carriers with habitual alcohol consumption, three out of six (50 %) *ALDH2*-deficient variant carriers without habitual alcohol consumption, and two out of five (40 %) *ALDH2*-deficient variant carriers with habitual alcohol consumption. Notably, no significant differences were observed between these groups (p = 0.95).

## Discussion

The present study demonstrated that the *ALDH2*-deficient variant itself was not inherently a risk factor for AF recurrence following catheter ablation. However, individuals with this variant had an elevated risk of AF recurrence, particularly when accompanied with habitual alcohol consumption, even after catheter ablation. Moreover, the risk of AF recurrence in *ALDH2*-deficient variant carriers increased depending on the amount of alcohol consumed. In contrast, the risk of AF recurrence in moderate and heavy drinkers among *ALDH2* wild-type carriers appeared to be similar.

Comparisons of the results of AF catheter ablation between Asian and Caucasian populations showed almost identical success rates (7,8). However, there are differences in the characteristics of each population, particularly in the proportion of obese individuals. In addition to habitual alcohol consumption, obesity is closely linked to AF recurrence following catheter ablation (14,15). In a multicenter cohort study, the mean BMI of Japanese patients was approximately 24 kg/m (8), whereas that of the Caucasian population was 30 kg/m (7). While the recurrence rate in studies on Caucasian populations is expected to be higher than that in Japanese populations, we found that their AF recurrence rates were similar. Studies have reported that the presence of *ALDH2*-deficient variant carriers is rare in Caucasians, whereas it is more prevalent, ranging 30 to 40 % in East-Asian populations (22,23). These findings and the results of this present study suggest that this discrepancy may be attributed to the *ALDH2* genotypes.

In the present study, the weekly volume of alcohol consumption among *ALDH2* wild-type and *ALDH2*-deficient variant carriers was 252 (range, 168-392) g/week and 224 (range, 98-315) g/week, respectively (p = 0.07). Moreover, while the low-voltage area in the left atrium of patients with habitual alcohol consumption was larger than that of those without habitual alcohol consumption, the low-voltage area of *ALDH2*-deficient variant carriers and *ALDH2* wild-type carriers with habitual alcohol consumption did not significantly differ. The volume of alcohol consumption and the low voltage area in the left atrium of the patients did not show significant differences between the *ALDH2* genotypes. However, the recurrence rate in *ALDH2*-deficient variant carriers was significantly higher than that in *ALDH2* wild-type carriers. These results suggest that alcohol itself may not be the primary factor influencing AF recurrence; rather, acetaldehyde, the ultimate alcohol metabolite processed by ALDH2, might play a more crucial role in AF recurrence. Additionally, overdrive pacing failed to induce triggered activity in the presence of ethanol; however, the triggered activity was successfully induced in the presence of acetaldehyde (28). Therefore, the mechanism of AF recurrence, even performed catheter ablation, may be triggered activity with the AF substrate, representing a wide low-voltage area. The fact that PV reconnection was not significantly different between the four patient groups in the present study does not contradict this speculation.

To reduce AF recurrence following catheter ablation, it may be important to identify whether an individual is a carriers of the *ALDH2*-deficient variant, especially in the Asian population. This is significant because the prevalence of *ALDH2*-deficient variant carriers is notably higher in Asian populations than in other populations (22), (23). Previous research has shown that recognizing the symptoms of alcohol flushing syndrome can serve as a predictor to differentiate between *ALDH2* wild-type and *ALDH2*-deficient variants. As a result, providing guidance on alcohol abstinence may be important, especially in patients exhibiting alcohol flushing syndrome, even after catheter ablation is performed. In fact, although it needs to be investigated further, patients with alcohol flushing syndrome attributed to *ALDH2*-deficient tend to stop alcohol consumption after receiving alcohol abstinence guidance more than *ALDH2*-wild type carriers.

Regarding the relationship between *ALDH2*-deficient variant and other disease, some reports have suggested that *ALDH2*-deficient variant contributes to coronary spastic angina (21) and upper aerodigestive tract cancer (29) (30) when accompanied with habitual alcohol consumption. The mechanism of this phenomenon has been clarified to the accumulation of acetaldehyde due to the slow metabolize of alcohol (30). And this mechanism is similar to the speculation of this present study. Therefore, identify the *ALDH2*-genotype and education of *ALDH2* genotype-alcohol relationship might lead to prognostic improvement owing to reducing not only AF but other cardiovascular disease and upper aerodigestive tract cancer. From above findings, the present study may contribute to the development of personalized therapeutic approaches for *ALDH2*-deficient variant carriers.

### Study limitations

This study has a few limitations. It has been demonstrated that sleep apnea syndrome is closely associated with AF recurrence following catheter ablation. However, in this study, we could not evaluate its contribution to AF recurrence owing to the limited number of patients diagnosed with sleep apnea syndrome (six patients). To better assess the impact of sleep apnea syndrome on AF recurrence, we would have been required to conduct additional polysomnography examinations on the remaining patients, potentially leading to an increase in the number of identified cases. Nevertheless, it is important to acknowledge that with more comprehensive data, findings regarding the relationship between sleep apnea syndrome and AF recurrence may evolve.

As described above, the *ALDH2*-deficient variant is comprised of heterozygous-deficient (*1/*2) and homozygous-deficient (*2/*2) alleles. While categorization was possible, the analysis could not be applied due to insufficient previous data, particularly *ALDH2*\*2/*2 carriers in related to AF recurrence. Therefore, the analysis, where *ALDH2* genotypes were categorized into *ALDH2* wild-type, *ALDH2*\*1/*2, and *ALDH2*\*2/*2 carriers, is described in the supplemental materials.

## Conclusion

While the *ALDH2*-deficient variant itself was not found to be directly associated with AF recurrence, it emerged as a strong risk factor when accompanied with habitual alcohol consumption. Thus, abstaining from alcohol consumption may play a crucial role in reducing the risk of AF recurrence, especially for patients with alcohol flushing syndrome, which serves as a predictor for *ALDH2*-deficient variant carriers, even after catheter ablation has been performed. In addition, these results may contribute to the development of personalized therapeutic approaches for patients undergoing catheter ablation.

## Data Availability

The datasets used and analyzed during the current study available from the corresponding author on reasonable request.

## Acknowledgements

The author would like to thank Mr. Shogo Tsuda, Yoshihisa Kiyota, and Yoshitaka Sakata for technical support.

## Non-standard Abbreviations and Acronyms

ALDH2: Aldehyde dehydrogenase 2

AAD: Antiarrhythmic drug

AF: Atrial fibrillation

CI: Confidence interval

HR: Hazard ratio

PV: Pulmonary vein

## Central Illustration

Relationship between *ALDH2* genotypes and atrial fibrillation recurrence following catheter ablation. AF, atrial fibrillation; ALDH2, aldehyde dehydrogenase 2; CI, confidence interval;

